# Early detection and treatment of postpartum haemorrhage: a cost-effectiveness analysis of the E-MOTIVE trial

**DOI:** 10.1101/2024.01.11.23300121

**Authors:** E.V. Williams, I. Goranitis, R. Oppong, S.J. Perry, A. Devall, J.T. Martin, K-M. Mammoliti, L. Beeson, K.N. Sindhu, H. Galadanci, F. Alwy Al‑beity, Z. Qureshi, G.J. Hofmeyr, N. Moran, S. Fawcus, S. Mandondo, L. Middleton, K. Hemming, O. Oladapo, I. Gallos, A. Coomarasamy, T.E. Roberts

## Abstract

**Background:** Timely detection and treatment of postpartum haemorrhage (PPH) are crucial to prevent complications or death. A calibrated blood-collection drape can help provide objective, accurate, and early diagnosis of PPH and a treatment bundle can address delays or inconsistencies in the use of effective interventions.

**Methods:** We conducted an incremental cost-effectiveness analysis alongside the E-MOTIVE trial, an international, parallel cluster-randomised trial with a baseline control phase, designed to assess a multi-component intervention for PPH in patients having vaginal delivery. We compared the E-MOTIVE intervention, which included a calibrated blood-collection drape for early detection of PPH and a bundle of first-response treatments (uterine massage, oxytocic drugs, tranexamic acid, intravenous fluids, examination, and escalation), with usual care. We used multilevel modelling to estimate incremental cost-effectiveness ratios from the perspective of the public healthcare system for outcomes of cost per case of severe PPH (blood loss ≥1000 mL) prevented and cost per disability-adjusted life-year (DALY) averted.

**Results:** A total of 80 secondary-level hospitals across Kenya, Nigeria, South Africa, and Tanzania, in which 210,132 patients underwent vaginal delivery, were randomly assigned to the E-MOTIVE group or the usual-care group. Among hospitals and patients with data, severe PPH was diagnosed in 1.6% of patients in the E-MOTIVE group and 4.3% of patients in the usual-care group (risk difference, -2.6%; 95% CI -3.1% to -2.1%). Mean DALYs per patient were lower for the E-MOTIVE group (-0.0027; 95% CI -0.0081 to 0.0029) whilst mean costs per patient were slightly higher compared with the usual-care group (0.30 USD; 95% CI -2.31 to 2.78). The E-MOTIVE intervention was deemed cost-effective at contemporary willingness-to-pay thresholds and remained cost-effective across the full range of sensitivity and country-level analyses.

**Interpretation:** Use of a calibrated blood-collection drape for early detection of PPH and bundled first-response treatment is cost-effective and should be perceived by decision makers as a worthwhile use of healthcare budgets.

**Funding:** Bill & Melinda Gates Foundation (NCT04341662).

## Introduction

Postpartum haemorrhage (PPH), defined as blood loss ≥ 500 mL from the genital tract after childbirth, is the leading cause of maternal death worldwide, accounting for approximately 27% of maternal deaths.^1,2^ PPH is a major concern in low- and middle-income countries (LMICs), where PPH-associated mortality is disproportionately high.^3^ PPH is associated with considerable economic burden: recent estimates from four LMICs suggest the costs of direct hospital care for women with PPH can be up to 2.8 times higher than for a birth without PPH.^4^ In addition, the immediate and long-term economic consequences of maternal mortality incurred by households can be substantial.^5-7^

The World Health Organization (WHO) has published and updated several evidence-informed recommendations for the prevention and treatment of PPH.^8,9^ However, adherence to these recommendations in many low-resource settings is limited by numerous challenges. Firstly, PPH is often undetected or detected late; consequently life-saving treatment is not promptly initiated. The current usual practice of blood-loss assessment is visual estimation, which is widely recognised as inaccurate and typically leads to underestimation of blood loss.^10^ An additional challenge is delayed or inconsistent use of effective interventions for management of PPH. Treatments for PPH are often administered sequentially; healthcare providers wait to observe the effects of one intervention before administering another intervention.^11^ However, PPH is a time-critical condition, and such delays can result in loss of life. Some cost-effective interventions may not be used at all: evidence from hospitals in Kenya, Nigeria, South African and Tanzania showed that tranexamic acid (TXA) was administered late and mostly as a last resort for women requiring surgery due to PPH.^12^ Furthermore, despite the availability of clear recommendations regarding PPH and their wide dissemination, uptake at the point of care remains low.^13^ An underpinning factor to some of the challenges relates to limited resources, therefore it is imperative to evaluate the resource implications of new interventions for managing PPH.

To address these challenges, the E-MOTIVE trial was funded by the Bill and Melinda Gates Foundation to assess a multicomponent intervention for detection and treatment of PPH in patients having vaginal delivery. The E-MOTIVE intervention consisted of a calibrated blood-collection drape for early detection of PPH and the WHO first-response bundle, which included uterine massage, oxytocic drugs, TXA, intravenous (IV) fluids, and a process for examination and escalation. The clinical effectiveness of the E-MOTIVE intervention has already been reported.^14^ Here we report the economic evaluation conducted alongside the E-MOTIVE trial, an integral component of the E-MOTIVE project, which aimed to assess the cost-effectiveness of the E-MOTIVE intervention compared with usual care. The economic evaluation, which was carried out from a healthcare system perspective, was based on the outcomes of cost per case of severe PPH prevented (blood loss, ≥1000 mL), and cost per disability-adjusted life-year (DALY) averted.

## Methods

The economic evaluation assessed the cost-effectiveness of the E-MOTIVE intervention compared with usual care. Data on health outcomes and resource use were collected prospectively during the E-MOTIVE trial. Two main analyses were conducted from the healthcare system perspective, based on two outcomes: cost per case of severe PPH prevented and cost per DALY averted. We used multilevel modelling to estimate incremental costs and outcomes across the whole sample, and present results using incremental cost-effectiveness ratios (ICERs). We conducted deterministic sensitivity analysis on cost inputs for the base-case and probabilistic sensitivity analyses to quantify decision uncertainty. An additional analysis was conducted from the perspective of each participating country, based on whole trial data, to present country-level estimates to provide local context for decision makers.

### Study design and participants

The E-MOTIVE trial was an international, parallel cluster-randomised trial that included a baseline control phase. Detailed information about the trial design and participants is published elsewhere.^14^ Briefly, between August and October 2021, 80 secondary-level hospitals (14 in Kenya, 38 in Nigeria, 14 in South Africa, and 14 in Tanzania) entered a 7-month baseline period during which they provided usual care for PPH in patients having vaginal delivery. After this 7-month baseline period, hospitals were randomly assigned, in a 1:1 ratio, to continue providing usual care or to receive the E-MOTIVE intervention for 7 months, with a 2-month ‘transition phase’ to allow hospitals to adapt clinical practices for intervention delivery. Two hospitals in Tanzania did not receive the assigned intervention due to participation in a conflicting program, therefore were not included in the analysis.

Inclusion and exclusion criteria are detailed elsewhere.^14^ Approval was granted by the University of Birmingham, the Ethics Review Committee of the World Health Organization (WHO) (for the formative phase), and the relevant ethics and regulatory review committees in each participating country.

### Intervention

The E-MOTIVE intervention consisted of a blood-collection drape, with calibrated lines to measure blood-loss volume, for early detection of PPH and the WHO first-response treatment bundle, which included uterine massage, oxytocic drugs, TXA, IV fluids, and a process for examination and escalation (figure 1). Detailed information on the intervention is published elsewhere.^14^

**Figure 1.**
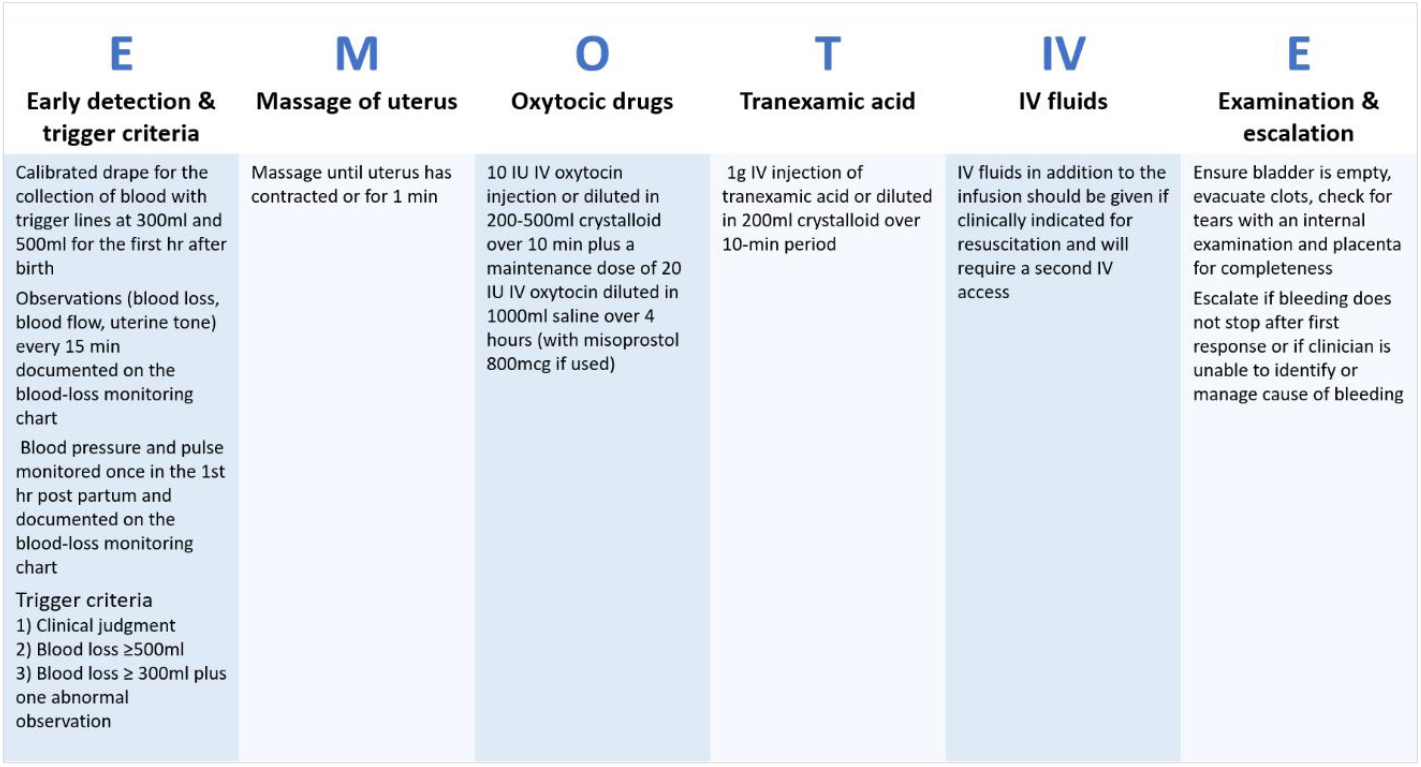
E-MOTIVE Intervention.

In usual care, blood loss is estimated visually, and first-response treatment may consist of some or all of the components of the WHO first-response bundle. These were typically administered sequentially, with oxytocic drugs given as first-line treatment and TXA reserved for refractory bleeding. Uncalibrated drapes, without alert or action lines, were used in the usual-care group hospitals to quantify blood loss for the purpose of the trial. We applied established dosage regimens for usual care, consistent with the E-MOTIVE group.

### Effectiveness outcomes

We estimated cost-effectiveness based on outcomes of severe PPH prevented and disability-adjusted life-years (DALYs) averted.

#### Severe PPH prevented

Severe PPH, defined as blood loss of at least 1000 mL, was measured at one hour and if there was continued bleeding, for up to two hours post-partum. Blood loss was objectively measured with the use of a blood-collection drape. Calibrated drapes were used in the hospitals in the E-MOTIVE group to enable early and accurate diagnosis of PPH and to obtain data on blood loss. Uncalibrated drapes were used in the hospitals in the usual-care group to obtain data on blood loss. Data on blood loss were source-verified by capturing a photograph of the drape with collected blood inside it, on a digital weighing scale, with the weight visible in the photograph. Only data that had been source-verified were used in the analysis.

#### DALYs averted

The DALY is a composite summary measure of disease burden that accounts for both mortality and non-fatal health consequences and is the preferred metric for economic evaluations to support resource allocation decisions in LMICs.^15^ DALYs were estimated for each treatment group based on non-fatal PPH events and maternal death from bleeding.

For non-fatal PPH events, years lived with disability (YLD) were estimated based on the magnitude of the disability and its duration. Disability weights for severe PPH (0.324 (≥ 1000 mL blood lost)) and less severe PPH (0.114 (< 1000 mL blood lost)) were drawn from the Global Burden of Disease (GBD) study.^16^ The duration of disability due to PPH (both severe and less severe) was considered to last for a postpartum period of six weeks. Given the trigger criterion of the E-MOTIVE intervention imposes a benefit on less-severe PPH it was imperative to include disability for less-severe PPH to ensure relevant effects were captured.

Years of life lost (YLL) for premature death due to bleeding were calculated using life expectancy of country-specific female populations drawn from GBD abridged life tables.^17^ YLL were calculated using a discount rate of 3%.

#### Resource use and costs

Resource use information was collected prospectively via case report forms (CRFs). Information was collected from the perspective of the healthcare system for calibrated drapes, uterotonic drugs, TXA, IV fluids, duration of hospitalisation, intensive care unit (ICU) admission, transfer to a higher-level facility, blood transfusions, postpartum laparotomy, hysterectomy, non-pneumatic anti-shock garments (NASGs), uterine balloon tamponades (UBTs) and bimanual compression. When necessary, data from an observational study conducted alongside the E-MOTIVE trial and expert clinical opinion from within the research study team supplemented CRF information.

Table A1 (supplementary appendix, p1) shows the unit costs used in the analysis. Calibrated blood-collection drape costs were obtained from Excellent Fixable Drapes in India, the manufacturer and supplier of the drapes used in the E-MOTIVE trial. We considered the price at which the drapes are currently being procured, 1.25 USD, in our base-case analysis. Costs of oxytocic drugs and TXA were obtained from a recent publication by the United States Agency for International Development (USAID) Global Health Supply Chain Program.^18^ Uterotonic drug costs were sourced from the United Nations Populations Fund (UNFPA) Product Catalogue, whilst the TXA costs reported were USAID wholesale prices. We obtained costs of IV fluids from the International Medical Product Price Guide (IMPPG), a recommended source of medication costs in LMIC settings.^19^ An adjustment of 25% was used to account for shipping, and handling charges, and internal distribution of traded goods.^20^

Country-specific unit cost estimates for bed days in secondary-level hospitals were obtained from the WHO-CHOICE initiative.^21,22^ Country-specific personnel costs were obtained from publicly available records regarding health sector pay, and personal communication with E-MOTIVE country trial management groups;^23^ costs from the latter were based on local government salaries. Conservative estimates of the lowest grade doctor who could attend a case of severe PPH were used. We used other secondary sources to estimate the cost of blood transfusions, additional treatment interventions, transfer to a higher-level facility and ICU admission.^4,24-28^

Due to a lack of cost data for postpartum laparotomy, we assumed a unit cost equivalent to 80% of a hysterectomy, based on expert clinical opinion from within the E-MOTIVE study team. Furthermore, we estimated unit costs for bimanual compression based on personnel requirements and procedure duration, and for UBT in Kenya, Nigeria, and Tanzania, we estimated costs considering materials and labour required for an improvised device. For the base case, we did not apply unit costs to activities perceived as a reprioritisation of existing staff time i.e., uterine massage and examination, as we assumed no additional resource was required. Additional details on costing assumptions are provided in the supplementary appendix (p2).

To standardise unit costs across countries where data were unavailable, a market basket approach was used, wherein an index table based on WHO-CHOICE estimates (supplementary appendix, p3) was used to indicate the relative mean cost of estimates for inpatient and outpatient health service delivery for each country-pair in the study.^20-22^ The market basket approach is an established costing method for the development of a complete set of country-specific unit cost data in the economic evaluation of multinational trials.^20^ All unit costs were adjusted to 2022 USD using average exchange rates and the average US inflation rate between the price base year used in individual studies and 2022, as recommended when there is a relatively high proportion of imported commodities in economic analyses.^29^ Given the short follow-up period of the trial, costs were not discounted.

### Statistical analysis

#### Main analysis

The economic evaluation comprised two main analyses: a cost-effectiveness analysis (CEA) based on the outcome of cost per case of severe PPH prevented and a cost-utility analysis (CUA) based on the outcome of cost per DALY averted. Both were carried out on an intention to treat basis and relied on complete case analysis wherein cases without source-verified blood loss data were excluded.

Following recommendations for the economic evaluation of cluster and multinational trials,^30,31^ we used multilevel modelling to estimate the difference in mean costs and outcomes between the E-MOTIVE and usual-care groups. Multilevel modelling accounts for unobserved cluster-specific effects on costs and outcomes and facilitates the estimation of cost-effectiveness across the whole sample.^32^ Consistent with the clinical analysis, we fitted generalised linear mixed models incorporating a constrained baseline analysis.^14^ For severe PPH, we used the binomial family and logit link, in addition to robust standard errors, followed by marginal standardisation to estimate risk difference. Differences in mean costs and DALYs were estimated using the Gaussian family and identity link, in combination with non-parametric permutation tests given the inherent skewness of such data. We included fixed effects for allocated exposure to E-MOTIVE, time period, country and covariates used in the randomisation method (number of vaginal births per hospital, the proportion of patients with a clinical primary-outcome event at each hospital, and the quality of oxytocin at each hospital during the baseline phase). We adjusted for clustering using random cluster and cluster-by-period effects.

Model estimates of the difference in costs and outcomes were used to derive an incremental cost per case of severe PPH prevented and an incremental cost per DALY averted. For the CUA, we used two thresholds to judge the cost-effectiveness of the E-MOTIVE intervention (table 1): a weighted threshold based on the WHO recommended threshold for a ‘highly cost-effective’ intervention of the countries’ per capita gross domestic product (GDP) and a weighted threshold based on recently advocated opportunity-cost based thresholds put forward by Woods and colleagues,^33,34^ equivalent to 51% GDP per capita for Kenya, Nigeria and Tanzania, and 71% GDP per capita for South Africa.

**Table 1.** Willingness-to-pay (USD) estimates for a disability-adjusted life-year averted in the participating countries.

| Recommendation | Kenya | Nigeria | South Africa | Tanzania | Weighted mean |
| --- | --- | --- | --- | --- | --- |
| WHO – per capita GDP | 2099 | 2184 | 6776 | 1192 | 2816 |
| Estimate based on Woods *et al.* (2016) | 1071 | 1114 | 4811 | 608 | 1690 |

Weighted cost-effectiveness thresholds are based on the proportion of participants from each country out of trial sample size. GDP per data obtained from the World Bank data set.<sup>35</sup>

#### Sensitivity analysis

We conducted sensitivity analyses to quantify the uncertainty relating to key assumptions and sampling variations. To characterise the inherent uncertainty around incremental cost-effectiveness estimates, we used non-parametric clustered bootstrapping with multilevel models to generate 1000 paired estimates of incremental mean total costs and DALYs. These estimates were used to construct a cost-effectiveness acceptability curve, which shows the probability that the E-MOTIVE intervention is cost-effective across a range of willingness-to-pay (WTP) threshold values per additional DALY averted. We also conducted deterministic sensitivity analyses on input parameters for the base-case analysis, including varying the device cost of the calibrated drapes to 1 USD, 0.75 USD and 0.50 per unit (current prices) respectively, considering potential cost decreases with expanded production.

#### Country-level analysis

To provide local context for decision makers, we estimated the cost-effectiveness of the E-MOTIVE intervention from the perspective of each participating country using four fully pooled, single-country costing CUAs. Clinical outcome and utilisation data from all participating countries were pooled, and country-specific unit costs and life-expectancy data were applied to all patients in the trial. The country-level analyses were adjusted analogously to the main analyses. Model estimates of differences in cost and DALYs were used to derive ICERs, which were judged against the country-specific thresholds reported in Table 1.

All analyses were carried out using Stata, version 17.1 (StataCorp).

## Results

Data for analysis were obtained from 78 secondary-level hospitals (from 14 in Kenya, 38 in Nigeria, 14 in South Africa, and 12 in Tanzania), with a total of 210,132 patients (110,473 in the baseline phase and 99,659 in the implementation phase) giving birth vaginally in the hospitals during the trial period. Source-verified data regarding blood loss were available for 206,455 patients (107,733 in the baseline phase and 98,722 in the implementation phase; 98% follow-up).

Severe PPH was diagnosed in 786 of 48,678 patients (1.6%) in the E-MOTIVE group and in 2129 of 50,043 (4.3%) in the usual-care group (adjusted risk difference, -2.6% (95% CI -3.1% to -2.1%; table 2). In the E-MOTIVE group the mean DALYs per patient was 0.00767 (SD 0.394) and in the usual-care group the mean DALYs per patient was 0.01158 (SD 0.454). The adjusted DALY difference between E-MOTIVE and usual care per patient was -0.00266 (95% CI -0.00814 to 0.00287, table 2).

**Table 2.** Mean per-patient total costs (2022 USD) and DALYs, risk of severe PPH and ICERs.

|  | **E-MOTIVE**<br>**(N = 48,678)** | **Usual care**<br>**(N = 50,043)** | **Adjusted difference\*\***<br>**(95% CIs\*\*\*)** | **ICER (USD)** |
| --- | --- | --- | --- | --- |
| **Mean per-patient total cost (USD)** | 45.14<br>(107.93) | 43.19<br>(126.84) | 0.30<br>(-2.31 to 2.78) |  |
| **Mean per-patient DALYs** | 0.00767<br>(0.394) | 0.01158<br>(0.454) | -0.00266<br>(-0.00814 to 0.00287) | **113.91** |
| **Severe PPH\*** | 786<br>(1.6) | 2129<br>(4.3) | -2.6<br>(-3.1 to -2.1) | **11.83** |
| Values are mean (SD) or *number (percentage). Adjusted difference between severe PPH risks is presented in percentage points, and differences between mean values are presented in the unit of the values.<br>**Adjusted for number of vaginal births per hospital, time period, country, the proportion of patients with a clinical primary-outcome event at each hospital and the quality of oxytocin at each hospital during the baseline phase and for clustering using random cluster and cluster-by-period effects. Baseline data before implementation of the intervention (107,733 patients in 78 clusters) for the intervention and usual-care groups are as follows: for mean total cost (USD), 45.43 (134.05) in the E-MOTIVE group and 42.05 (145.37) in the usual-care group; for mean DALYs, 0.01037 (0.427) in the E-MOTIVE group and 0.01314 (0.490) in the usual-care group; for severe PPH, 1,920/50,720 (3.8) in the E-MOTIVE group and 2,535/57,010 (4.4) in the usual-care group.<br>*** For total costs and DALYs confidence intervals were constructed using non-parametric permutation tests, by finding the upper and lower boundaries of the intervention effect that would lead to a two-sided P value less than the 5% level (1000 replications). |  |  |  |  |

The resource utilisation per group is shown in the supplementary appendix (p3). Notably, administration of oxytocin, TXA and IV fluids - three core elements of the MOTIVE first-response bundle - was more common in the E-MOTIVE group despite lower rates of PPH (8.5% compared with 16.7% in the usual-care group). This can be explained by the improved detection of PPH facilitated by the use of a calibrated blood-collection drape and consequent triggering of the bundle. The usual-care group experienced higher numbers of blood transfusions, marginally longer hospitalisation, and greater need for additional treatment interventions. Also, significantly more severe PPH cases in the usual-care group necessitated additional time for physician attendance.

Detailed mean per-patient costs are shown in the supplementary appendix (p4). The total unadjusted mean per patient cost was 45.15 USD (SD 107.93) in the E-MOTIVE group and 43.19 USD (SD 126.84) in the usual-care group (table 2). The adjusted total cost difference was 0.30 USD (95% CI -2.31 to 2.78, table 2). The estimated ICERs (table 2) are therefore 11.83 USD per case of severe PPH averted and 113.91 USD per DALY averted. The ICER in terms of DALYs is below both the weighted GDP-based threshold (2816 USD) and opportunity-cost based threshold (1690 USD), suggesting the E-MOTIVE intervention is cost-effective. Figure 2 shows the probability of the E-MOTIVE intervention being cost-effective compared with usual care across a range of WTP thresholds per DALY averted. For thresholds of WTP per DALY averted greater than approximately 1500 USD, there is >80% probability that the E-MOTIVE intervention is cost-effective (figure 2).

**Figure 2.**
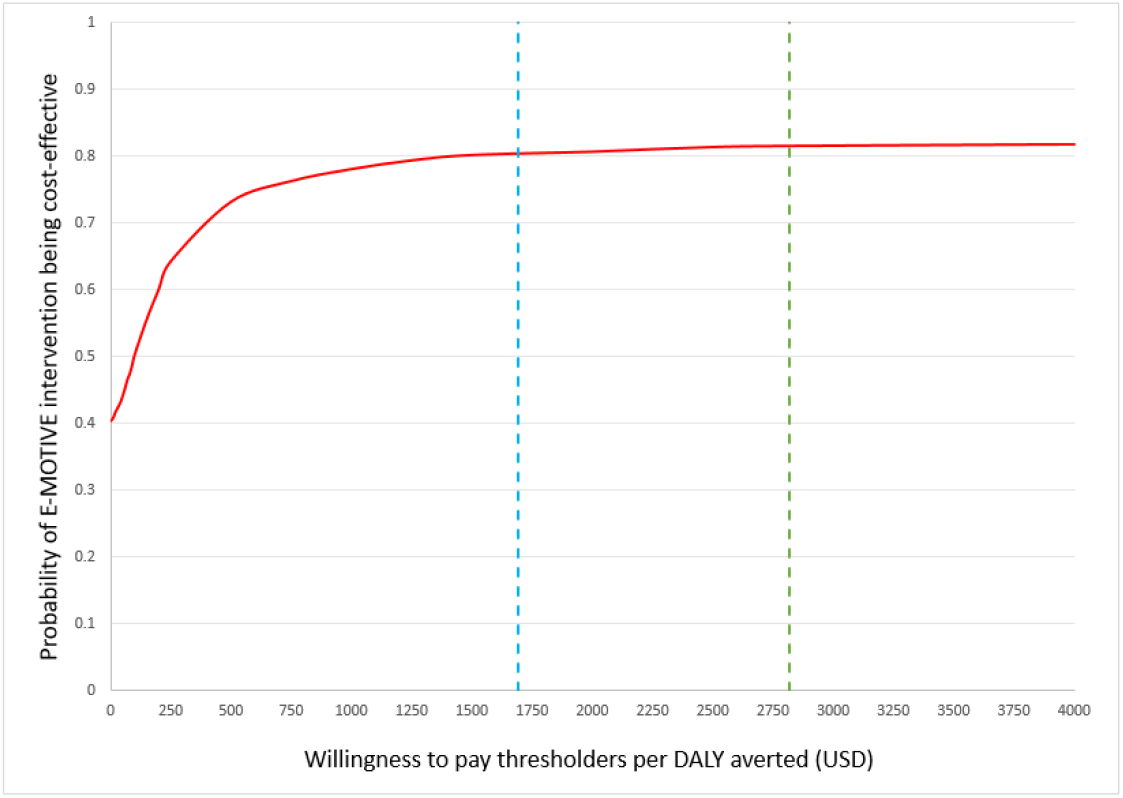
Cost-effectiveness acceptability curve indicating the probability of the E-MOTIVE intervention being cost-effective across different willingness-to-pay thresholds for a DALY averted. The dashed lines show the expected willingness-to-pay for a DALY averted, as estimated from WHO recommendations (green) and Woods and colleagues (blue).

For the sake of brevity, only variation in the cost of the calibrated drape is presented in table 3. If the device cost of the calibrated drape is reduced to 1 USD, the E-MOTIVE intervention becomes comparable in cost to usual care, whilst being more effective. Further reductions in the cost of the calibrated drape could potentially result in cost savings. Additional sensitivity analyses to test the assumptions were carried out but are not reported here as there was no substantial difference to the conclusion of the base-case results observed.

**Table 3.** Deterministic sensitivity analyses varying calibrated drape device cost.

| Sensitivity analysis | Mean total cost difference (USD)* | Mean DALY difference | ICER (USD) |
| --- | --- | --- | --- |
| Drape cost – 1 USD | -0.01 | 0.00266 | **Dominant\*\*** |
| Drape cost – 0.75 USD | -0.30 | 0.00266 | **Dominant\*\*** |
| Drape cost – 0.50 USD | -0.61 | 0.0026 | **Dominant\*\*** |

Device costs of calibrated blood-collection drapes are reported prior to adjustments to 2022 USD and for shipping, handling, and internal distribution.
\* Adjusted for number of vaginal births per hospital, time period, country, the proportion of patients with a clinical primary-outcome event at each hospital and the quality of oxytocin at each hospital during the baseline phase and for clustering using random cluster and cluster-by-period effects.
\*\* Dominance is based on point estimate only.

The mean costs, DALYs and ICERs from the country-level analyses are shown in table 4. Briefly, the E-MOTIVE intervention was judged to be cost-effective for each participating country when the ICERs were compared against both country-specific GDP-based WTP thresholds and opportunity-cost based thresholds (table 1). In South Africa, where the cost of calibrated drapes was lower relative to other resources, the E-MOTIVE intervention was less expensive and therefore the dominant intervention based on the point estimates.

**Table 4.** Country-level estimates of mean per-patient total costs (2022 USD), DALYs, and ICERs.

|  |  |  |  |  |
| --- | --- | --- | --- | --- |
| Country | E-MOTIVE | Usual Care | Adjusted Difference*<br>(95% CIs**) | ICER<br>(USD) |
| Kenya |  |  |  |  |
| Mean per-patient total cost (USD) | 14.86<br>(21.25) | 14.18<br>(23.58) | 1.08<br>(-0.68 to 2.75) | 402.51 |
| Mean per-patient DALYs | 0.00765<br>(0.39229) | 0.01157<br>(0.45407) | -0.00268<br>(-0.00817 to 0.00281) |  |
| Nigeria |  |  |  |  |
| Mean per-patient total cost (USD) | 22.20<br>(26.11) | 21.97<br>(29.87) | 0.66<br>(-1.99 to 3.24) | 248.57 |
| Mean per-patient DALYs | 0.00747<br>(0.39351) | 0.01157<br>(0.45407) | -0.00265<br>(-0.00814 to 0.00283) |  |
| South Africa |  |  |  |  |
| Mean per-patient total cost (USD) | 157.89<br>(198.30) | 168.99<br>(229.10) | -5.24<br>(-23.41 to 12.88) | Dominant*** |
| Mean per-patient DALYs | 0.00760<br>(0.38893) | 0.01148<br>(0.44881) | -0.00264<br>(-0.00806 to 0.00278) |  |
| Tanzania |  |  |  |  |
| Mean per-patient total cost (USD) | 11.54<br>(15.95) | 10.56<br>(17.26) | 1.26<br>(-0.01 to 2.58) | 473.79 |
| Mean per-patient DALYs | 0.00767<br>(0.3931) | 0.01158<br>(0.45407) | -0.00266<br>(-0.00814 to 0.00284) |  |
| Values are mean (SD). Adjusted differences in costs and DALYs were estimated by fully pooled, single-country costing analyses wherein clinical outcome and utilisation data from all participating countries were pooled, and country-specific unit costs and life-expectancy data were applied to all patients. |  |  |  |  |
| *Adjusted for number of vaginal births per hospital, time period, country, the proportion of patients with a clinical primary-outcome event at each hospital, and the quality of oxytocin at each hospital during the baseline phase and for clustering using random cluster and cluster-by-period effects. |  |  |  |  |
| **Confidence intervals were constructed using non-parametric permutation tests, by finding the upper and lower boundaries of the intervention effect that would lead to a two-sided P value less than the 5% level (1000 replications). |  |  |  |  |
| *** Dominance is based on point estimates. |  |  |  |  |

## Discussion

### Summary of main findings

This study assessed the cost-effectiveness of early detection of PPH using a calibrated drape and treatment using the WHO first-response treatment bundle, which included uterine massage, oxytocic drugs, TXA, IV fluids, and a process for examination and escalation, compared with usual care. The findings suggest that early detection of PPH using a calibrated blood-loss collection drape and treatment with the WHO first-response bundle is cost-effective compared with usual care. Our sensitivity analysis suggested that for WTP values above 1500 USD per DALY averted there is more than an 80% probability of the E-MOTIVE intervention being effective. Furthermore, deterministic sensitivity analysis showed that potential reductions in the cost of the calibrated blood-collection drape could lead to cost savings.

### Strengths and limitations

To our knowledge, this study is the first to assess the cost-effectiveness of early detection of PPH and the use of bundled treatment. The study benefitted from a large sample size recruited from 78 hospitals across four countries, broad inclusion criteria to capture all patients with vaginal births in the trial hospitals, and a wide range of primary data. However, the study is not without limitations. Owing to the pragmatic design of the trial, extensive bottom-up costing of all resource items was not conducted. This naturally increased the uncertainty around the unit cost estimates used in the analysis. However, when feasible, cost estimates were obtained from established sources and other secondary sources based on bottom-up costing. Some assumptions were required to estimate country-specific unit costs when these were not available. All assumptions were agreed prior to any analysis being undertaken and sensitivity analyses exploring their importance found they did not substantially impact the cost-effectiveness results.

Furthermore, PPH can involve considerable out-of-pocket expenses for women and their family members. Owing to the pragmatic design of the trial, these expenses were not captured. Also, evidence suggests maternal mortality is associated with substantial economic costs to society. Given there were fewer cases of severe PPH and less severe PPH in the E-MOTIVE group, and maternal deaths, though rare, were in the same direction – it is likely that under a societal perspective of analysis, E-MOTIVE would produce even more favourable cost-effectiveness results.

Finally, this analysis was conducted alongside a large international, cluster-randomised trial with a baseline control phase which presents complexities with respect to data analysis, for example, randomisation took place at the cluster level, but outcomes were measured at the level of the individual. This was addressed using methods to account for the hierarchical nature of the data, and the analysis adjusted for imbalances in outcomes during the baseline phase across trial groups. In addition, due to the substantial loss of power that would be experienced by analysing countries in isolation, country-specific cost-effectiveness analyses were not conducted. However, we assessed cost-effectiveness from the perspective of each participating country based on whole trial data. To this end, we conducted single-country costing CUAs in which clinical data from all participating countries were pooled, and country-specific unit costs and life-expectancy data were applied to all patients in the trial. Although not fully country-specific, we believe these estimates provide useful indicative information on cost-effectiveness for decision-makers, given the widespread occurrence of visual blood loss estimation, and delayed and inconsistent use of effective PPH interventions, such as TXA, across countries.

## Policy Implications

Our findings suggest that early detection of PPH and bundled treatment is cost-effective. Provision of calibrated blood-collection drapes and use of bundled first-response treatment can be considered a worthwhile use of constrained healthcare budgets. Guidelines on PPH management should be updated to reflect the efficacy evidence from the E-MOTIVE trial and findings of this cost-effectiveness analysis.

## Data Availability

Any data requests for the clinical data should be made to the corresponding author of the E-MOTIVE clinical paper. No additional data are available for this economic evaluation.

