## Supplementary Appendix for "Early detection and treatment of postpartum haemorrhage: a cost-effectiveness analysis of the E-MOTIVE trial"

**Table A1. Unit Costs**

| Item | Kenya | Nigeria | South Africa | Tanzania | Sources | Other Information |
|---|---|---|---|---|---|---|
| Calibrated blood-collection drape* | 1.52 | 1.52 | 1.52 | 1.52 | Personal communication with Excellent Fixable Drapes | Per unit |
| Oxytocin* | 1.25 | 1.25 | 1.25 | 1.25 | [18] | Per patient. Cost includes oxytocin for initial infusion 10IU (0.42 USD) and maintenance infusion 20IU (0.84 USD). |
| Tranexamic acid (TXA) * | 2.74 | 2.74 | 2.74 | 2.74 | [18] | Per 1g/10mL ampoule |
| Administration of TXA | 0.80 | 0.28 | - | 0.28 | Personal communication with country TMGs | Per procedure. Cost includes 10 minutes of midwife time for slow bolus injection. |
| Intravenous (IV) fluids* | 0.54 | 0.54 | 0.54 | 0.54 | [19] | Per 500mL. Cost per mL is used across volumes. |
| IV fluid giving set and cannula* | 0.38 | 0.38 | 0.38 | 0.38 | [26] | Per unit |
| Ergometrine* | 0.73 | 0.73 | 0.73 | 0.73 | [18] | Per 200 mcg/ml injection in 1mL ampoule |
| Misoprostol* | 1.25 | 1.25 | 1.25 | 1.25 | [18] | Per 800mcg (4 x 200mcg tablets) |
| Hysterectomy | 185.21 | 258.86 | 1339.49 | 138.91 | [24] [4] [25] | Per procedure |
| Laparotomy | 148.17 | 207.09 | 1071.59 | 111.13 | | Per procedure. Cost is 80% of postpartum hysterectomy cost |
| Non-pneumatic anti-shock garment (NASG)* | 1.27 | 1.17 | 1.54 | 1.17 | [26] | Per procedure, based on 72 uses of NASG. |
| Bimanual compression | 2.40 | 0.85 | 6.35 | 0.86 | [23] Personal communication with country TMGs | Per procedure, Assumption: 30 minutes of midwife time |
| Uterine balloon tamponade (UBT)* | 1.19 | 0.93 | 6.90 | 0.93 | [26] [27] | Per procedure. For Kenya, Nigeria, and Tanzania the cost includes the components of UBT device (condom, catheter and syringe) and 5 minutes of midwife time. For South Africa the cost includes Ellavi device. |
| Attending doctor for severe PPH | 1.54 | 0.79 | 4.27 | 0.72 | [23] Personal communication with country TMGs | Per case of severe PPH. Cost is 10 minutes of doctor time |
| Non-ICU hospitalisation | 7.62 | 13.99 | 110.27 | 5.61 | [22] | Per day in hospital. Cost estimates represent the hotel component of hospital costs, i.e., excluding the cost of drugs and diagnostic tests. |
| ICU admission | 57.43 | 100.50 | 717.86 | 43.07 | [28] | Per day in ICU |
| Transfer to higher level facility | 19.35 | 34.25 | 54.74 | 14.51 | [24,25] | Per event |
| Blood transfusion | 95.64 | 40.08 | 288.55 | 71.73 | [4] | Per procedure. Cost assumes 2 units of whole blood were required for blood transfusions. |
| Needles and syringe* | 0.05 | 0.05 | 0.05 | 0.05 | [26] | Per unit |

*Tradable goods were adjusted for shipping, handling, and internal distribution (+25%).

### Costing assumptions

Several simplifying assumptions were necessary to estimate unit costs in the analysis. These were based on expert clinical opinion from within the E-MOTIVE study team and findings from an observational study conducted alongside the trial. All assumptions were agreed prior to any analysis being undertaken.

- Staff time was only costed when additional labour was considered essential.
- For cases of *severe* PPH, it was assumed that a doctor would attend for 10 minutes. We considered the lowest appropriate grade doctor who could attend.
- To avoid zero costs, patients with a length of stay less than 24 hours were assigned the cost of a full day on the ward which can be considered as a proxy for the cost of delivery.
- Patients with an ICU length of stay less than 24 hours were assigned the cost of a full day in ICU.
- The duration of bimanual compression was assumed to be 30 minutes.
- Two units of whole blood were assumed to be required for those who needed blood transfusions.
- The cost of a cannula and an IV giving set were applied to half of the women who had IV PPH treatment, given observational findings showed that approximately 50% of patients had prior IV access.
- A one-off cost for syringes and needles for PPH treatment with oxytocin, TXA and ergometrine was applied.
- In the absence of robust costs for laparotomy across all countries, an assumed cost equivalent to 80% of a hysterectomy was applied.
- Uterine massage and examination of the genital tract were not assumed to bear an additional cost when delivered as part of the E-MOTIVE intervention. These aspects of the first-response treatment bundle were considered relevant to a reprioritisation of care and not additional care. It was expected to be delivered by the same attending midwife so extra resource would not be required.
- The costs of the uncalibrated drapes used by the usual-care group in the E-MOTIVE trial were not considered by this analysis as they were used for research purposes.

The importance of assumptions was explored using deterministic sensitivity analyses.

| Table A2. Relative cost indices of participating countries based on WHO-CHOICE estimates | | | | |
|---|---|---|---|---|
| **To country:** | **From country:** | | | |
| | Kenya | Nigeria | South Africa | Tanzania |
| Kenya | 1.00 | 0.57 | 0.08 | 1.34 |
| Nigeria | 1.77 | 1.00 | 0.14 | 2.37 |
| South Africa | 12.56 | 7.20 | 1.00 | 16.83 |
| Tanzania | 0.75 | 0.42 | 0.06 | 1.00 |

| Table A3. Resource use per group | | |
|---|---|---|
| | **E-MOTIVE (N = 48,678)** | **Usual care (N = 50,043)** |
| Uterine massage | 5,762 (11.84) | 4,085 (8.16) |
| Oxytocin use | 5,864 (12.05) | 4,283 (8.56) |
| Tranexamic acid use | 5,796 (11.91) | 1,965 (3.93) |
| Intravenous fluids use | 5,851 (12.02) | 4,163 (8.32) |
| Examination of the genital tract | 5,476 (11.25) | 3,581 (7.16) |
| Ergometrine use | 124 (0.25) | 75 (0.15) |
| Misoprostol use | 2,587 (5.31) | 2,705 (5.41) |
| Laparotomy | 1 (0.00) | 3 (0.01) |
| Hysterectomy | 11 (0.02) | 6 (0.01) |
| Non-pneumatic anti-shock garment (NASG) use | 89 (0.18) | 38 (0.08) |
| Bimanual compression | 136 (0.28) | 553 (1.11) |
| Uterine balloon tamponade use | 44 (0.09) | 57 (0.11) |
| Blood transfusion | 1,063 (2.18) | 1,286 (2.57) |
| Transfer to higher level facility | 82 (0.17) | 17 (0.03) |
| Intensive care unit (ICU) admissions | 7 (0.01) | 28 (0.06) |
| Duration of hospitalisation (days)* | 1.09 (1.78) | 1.14 (2.06) |
| Duration of ICU hospitalisation (days)* | 2.00 (2.45) | 1.57 (1.53) |

Values are number (percentage) or mean (SD)*

Baseline data before implementation of the intervention (107,733 patients in 78 clusters) for the E-MOTIVE and usual-care groups are as follows: *Uterine massage:* E-MOTIVE: 5,074 (10.00), usual care: 4,537 (7.96); *Oxytocin use:* E-MOTIVE: 5,740 (11.32), usual care: 4,063 (8.88); *Tranexamic acid use:* E-MOTIVE: 2,246 (4.43), usual care: 1,221 (2.14); *Intravenous fluid use:* E-MOTIVE: 5,289 (10.43), usual care: 4,458 (7.82); *Examination of the genital tract:* E-MOTIVE: 4,007 (7.90), usual care: 3,786 (6.64); *Ergometrine use:* E-MOTIVE: 131 (0.26), usual care: 162 (0.28); *Misoprostol use:* E-MOTIVE: 3,286 (6.48), usual care: 3,234 (5.67); *Laparotomy:* E-MOTIVE: 3 (0.01), usual care: 3 (0.01); *Hysterectomy:* E-MOTIVE: 7 (0.01), usual care: 8 (0.01); *NASG:* E-MOTIVE: 72 (0.14), usual care: 50 (0.09); *Bimanual compression:* E-MOTIVE: 303 (0.60), usual care: 438 (0.77); *Uterine balloon tamponade:* E-MOTIVE: 23 (0.05), usual care: 56 (0.11); *Blood transfusion:* E-MOTIVE: 1,474 (2.91), usual care: 1,652 (2.90); *Transfer to higher level facility:* E-MOTIVE: 74 (0.15), usual care: 23 (0.04); *ICU admissions:* E-MOTIVE: 13 (0.03), usual care: 39 (0.07); *Duration of hospitalisation (days)**: E-MOTIVE: 1.06 (2.31), usual care: 1.05 (1.79); *Duration of ICU hospitalisation (days)**: E-MOTIVE: 2.00 (1.96), usual care: 2.02 (3.09).

| Table A4. Mean per-patient costs (2022 USD) | | | | |
|---|---|---|---|---|
| | **E-MOTIVE (N= 48,678)** | **Usual care (N = 50,043)** | **Adjusted Difference*** | **95% CIs**** |
| Calibrated blood-collection drape | 1.518 (0.000) | 0.000 (0.000) | 1.518 | - |
| Oxytocin | 0.151 (0.408) | 0.111 (0.357) | 0.034 | 0.007 to 0.068 |
| Tranexamic acid use | 0.367 (1.00) | 0.126 (0.624) | 0.252 | 0.199 to 0.305 |
| Intravenous fluids use | 0.216 (0.582) | 0.162 (0.512) | 0.047 | 0.007 to 0.087 |
| Misoprostol | 0.066 (0.280) | 0.068 (0.282) | 0.004 | -0.019 to 0.026 |
| Ergometrine | 0.002 (0.037) | 0.001 (0.026) | 0.002 | 0.000 to 0.003 |
| Needles and syringes | 0.006 (0.016) | 0.005 (0.014) | 0.001 | 0.000 to 0.002 |
| Laparotomy | 0.003 (0.672) | 0.009 (1.241) | -0.008 | -0.019 to 0.005 |
| Hysterectomy | 0.117 (10.875) | 0.025 (2.333) | 0.101 | -0.030 to 0.228 |
| Non-pneumatic anti shock garment (NASG) | 0.002 (0.051) | 0.001 (0.035) | 0.001 | -0.001 to 0.003 |
| Bimanual compression | 0.003 (0.093) | 0.013 (0.178) | -0.004 | -0.012 to 0.004 |
| Uterine balloon tamponade (UBT) | 0.002 (0.076) | 0.001 (0.055) | 0.000 | -0.001 to 0.002 |
| Blood transfusion | 1.348 (12.710) | 1.899 (16.940) | -0.088 | -0.585 to 0.439 |
| Non-ICU hospitalisation | 41.175 (104.015) | 40.526 (118.889) | -1.718 | -3.823 to 0.396 |
| ICU admission | 0.037 (3.996) | 0.439 (31.238) | 0.194 | -0.196 to 0.689 |
| Transfer to higher level facility | 0.089 (2.2017) | 0.011 (0.707) | 0.024 | -0.001 to 0.047 |
| Severe postpartum haemorrhage (doctor time) | 0.034 (0.334) | 0.0622 (0.3949) | -0.027 | -0.035 to -0.018 |
| **Mean total cost (USD)** | **45.135 (107.932)** | **43.189 (126.844)** | 0.302 | -2.312 to 2.783 |

Values are mean (SD).
*Adjusted for number of vaginal births per hospital, time period, country, the proportion of patients with a clinical primary-outcome event at each hospital and the quality of oxytocin at each hospital during the baseline phase and for clustering using random cluster and cluster-by-time effects.
** Confidence intervals were constructed using non-parametric permutation tests, by finding the upper and lower boundaries of the intervention effect that would lead to a two-sided P value less than the 5% level (1000 replications).
